# Establishing the relationships between adiposity and reproductive factors: a multivariable Mendelian randomization analysis

**DOI:** 10.1101/2023.03.03.23286615

**Authors:** Claire Prince, Laura D Howe, Gemma C Sharp, Abigail Fraser, Rebecca C Richmond

## Abstract

**Background:** Few studies have investigated associations between adiposity and reproductive factors using causal methods, both of which have a number of consequences on disease. Here we assess whether adiposity at different points in the lifecourse affects reproductive factors differently and independently, and the plausibility of the impact of reproductive factors on adiposity.

**Methods:** We used genetic data from UK Biobank and other consortia for eight reproductive factors: age at menarche, age at menopause, age at first birth, age at last birth, number of births, being parous, age first had sexual intercourse and lifetime number of sexual partners, and two adiposity traits: childhood body size and adulthood body mass index (BMI). We applied multivariable mendelian randomization to account for genetic correlation and estimate causal effects of childhood and adulthood adiposity, independently of each other, on reproductive factors. Additionally, we estimated the effects of reproductive factors, independently of other relevant reproductive factors, on adulthood adiposity.

**Findings:** We found a higher childhood body size leads to an earlier age at menarche, which in turn leads to higher adulthood BMI. Furthermore, we find contrasting and independent effects of childhood body size and adulthood BMI on age at first birth (Beta 0.22 SD (95% confidence interval:0.14,0.31) vs -2.49 (−2.93,-2.06) per 1 SD increase), age at last birth (0.13 (0.06,0.21) vs -1.86 (−2.23,-1.48) per 1 SD increase), age at menopause (0.17 (0.09,0.25) vs -0.99 (−1.39,-0.59) per 1 SD increase), and likelihood of having children (Odds ratio 0.97 (0.95,1.00) vs 1.20 (1.06,1.37) per 1 SD increase).

**Conclusions:** We highlight the importance of untangling the effects of exposures at different timepoints across the lifecourse, as demonstrated with adiposity, where accounting for measures at one point in the lifecourse can alter the direction and magnitude of effects at another time point and should therefore be considered in further studies.

## Introduction

Multiple observational studies have shown associations between women’s reproductive factors including age at menarche (AAM),^(1)^ age at first birth (AFB),^(1,2)^ number of births,^(1,3)^, age at menopause (AMP),^(4-7)^ and adiposity at different stages of the lifecourse. There remain a number of areas of active research: whether adiposity at difference points in the lifecourse affects reproductive factors, i.e., adiposity experienced in childhood and adulthood; whether any effects of adiposity at one point in the lifecourse, e.g., childhood, on reproductive factors is independent of adiposity at other points in the lifecourse e.g., adulthood; whether it is also plausible that reproductive factors affect adiposity, e.g., menopausal stage affecting concurrent and post-menopausal adiposity.^(8)^

Few studies have used causal methods to investigate the associations between adiposity and reproductive factors, and particularly considering time-varying adiposity.^(9)^ Mendelian Randomization (MR) is a method that can avoid problems of confounding and reverse causality, allowing the assessment of causality by using genetic variants associated with an exposure of interest as instrumental variables.^(10)^

Two MR studies: one using the UK Biobank study and the other the Avon Longitudinal Study of Parents and Children, with replication in the Genetic Investigation of Anthropometric Traits (GIANT) consortium, have suggested that earlier AAM causes higher adulthood BMI.^(11,12)^ However, the effect identified in the second study attenuated when adjusted for childhood BMI.^(12)^ Others have identified evidence to suggest a strong causal effect of higher childhood BMI on the risk of early menarche (i.e. a potential bidirectional relationship).^(13)^ An inverse relationship between adulthood BMI and AMP (mean age: 49.8 years, standard deviation (SD): 5.1) was found in an MR study using the UK Biobank, with replication in the ReproGen consortium.^(14)^ However, there have been limited MR studies investigating potential bidirectional causal relationships between adiposity and reproductive factors other than AAM and AMP.

We aimed to estimate the causal effects of childhood and adulthood adiposity, independently of each other, on a range of women’s reproductive factors. Additionally, we aimed to investigate the potential effects of reproductive factors on adulthood adiposity, independently of other reproductive factors, as we have shown reproductive factors to be genetically correlated with each other.^(15)^

## Methods

### UK Biobank

The UK Biobank study is a large population-based cohort of 502,682 individuals who were recruited at ages 37–73 years across the UK between 2006 and 2010. The study includes extensive health and lifestyle questionnaire data, physical measures, and biological samples from which genetic data has been generated. The study protocol is available online, and more details have been published elsewhere.^(16)^ At recruitment the participants gave informed consent to participate and be followed up.

### Reproductive factors

The reproductive factors investigated in this study were: AAM, AMP, age at first live birth, age at last live birth, number of live births, age first had sexual intercourse (AFS), lifetime number of sexual partners (at the time of assessment) and parous status (ever/never given birth at the time of assessment). Age at first live birth, age at last live birth and number of live births will hereafter be referred to as age at first birth, age at last birth and number of births. In UK Biobank these reproductive factors were derived from questionnaire responses at the baseline assessment; further details can be found in the **Supplementary Note**.

### Adiposity measures

We investigated adiposity in adulthood using BMI which was derived from height and weight measured during the initial UK Biobank Assessment Centre visit. In addition, we assessed adiposity in childhood using the comparative body size measure obtained from the baseline questionnaire in UK Biobank. Participants were asked, ““When you were 10 years old, compared to average would you describe yourself as:”, and were given the options: “Thinner”, “Plumper” and “About average”. The genetic score of this measure has been validated as a marker of childhood adiposity by previous studies in the Trøndelag Health Study,^(17)^ The Young Finn Study,^(18)^ and the Avon Longitudinal Study of Parents and Children,^(19)^ and the polygenic score for childhood body size in UK Biobank was more correlated with childhood obesity in an independent sample compared to the adulthood BMI genome-wide association studies (GWAS).^(20)^ In addition genetic risk scores for childhood body size are more strongly associated with fat mass compared to lean mass.^(21)^

### GWAS

To identify genetic variants robustly related to each of the reproductive factors, we performed GWAS for each reproductive factor among women in the UK Biobank. Each GWAS was performed using the Medical Research Council (MRC) Integrative Epidemiology Unit (IEU) UK Biobank GWAS pipeline.^(22,23)^ Further details can be found in the supplementary note.

We obtained female-only GWAS summary statistics for childhood body size and adulthood BMI from Richardson et al. 2020,^(19)^ where they performed GWAS using a similar approach (supplementary note).

### Univariable Mendelian randomization

We conducted MR analysis using the “TwoSampleMR” R package.^(23)^ The inverse variance weighted (IVW) method was used in the primary analysis to assess the causal relationships.

First, we assessed the causal effect of childhood body size on the eight reproductive factors. We then investigated the effects of adulthood BMI on seven of these reproductive factors, excluding AAM which precedes adulthood. Finally, we assessed the effect of each of the eight reproductive factors on adulthood BMI. Since all of the reproductive events occur after childhood, the effect of these on childhood body size was not considered.

The series of univariable MR (UVMR) analysis performed are shown in Table S1. GWAS estimates were standardized (mean = 0 and SD = 1) prior to performing MR.

Further details can be found in the supplementary note.

### Evaluating the impact of sample overlap, winner’s curse, and weak instruments

We used MRlap, a method which is robust to bias introduced by sample overlap, winner’s curse and weak instruments.^(24)^ This method only works in a univariable setting. MRlap was performed using the UK Biobank GWAS summary statistics for reproductive factors and adiposity measures, consistent with our primary univariable analysis.

### Multivariable Mendelian randomization

We performed multivariable MR (MVMR) analyses, an extension of MR,^(25,26)^ to estimate the direct effects of each reproductive factor, and adiposity measure by accounting for the genetic correlation between reproductive factors, and between adiposity measures in childhood and adulthood. Further details can be found in the supplementary note.

These analyses used the “MVMR” R package,^(27)^ to estimate the direct effects of childhood body size and adulthood BMI, mutually adjusting for the other adiposity measurement. This mutual adjustment aimed to account for genetic correlation between childhood body size and adulthood BMI.

Finally in the analyses evaluating the direct effect of each of the eight reproductive factors of interest on adulthood BMI, we adjusted for other reproductive factors in turn. The reproductive factors adjusted for were chosen based on previously identified causal relations between reproductive factors.^(15)^ The exposure, outcome and adjustment variables included in each MVMR model are shown in Table S2.

### Evaluating Mendelian randomization assumptions

We evaluated instrument strength, heterogeneity, pleiotropy and intended causal direction for the UVMR and MVMR model. Further details can be found in the supplementary note.

### Replication analyses

We performed replication analyses using GWAS summary statistics from samples with little or no overlap with UK Biobank. This allows us to evaluate the robustness of our results in both the UVMR and MVMR models. Replication GWAS summary statistics were obtained from the Early Growth Genetics (EGG) consortium, GIANT consortium, ReproGen and Social Science Genetic Association Consortium (SSGAC). All replication GWAS summary statistics were female-only other than those from the EGG consortium which were sex-combined.

Further details can be found in the supplementary note and Table S3.

## Results

### UK Biobank

273,238 women from UK Biobank were included. The mean age at assessment was 56 years (SD=8), further sample characteristics are shown in Table 1.

**Table 1.**
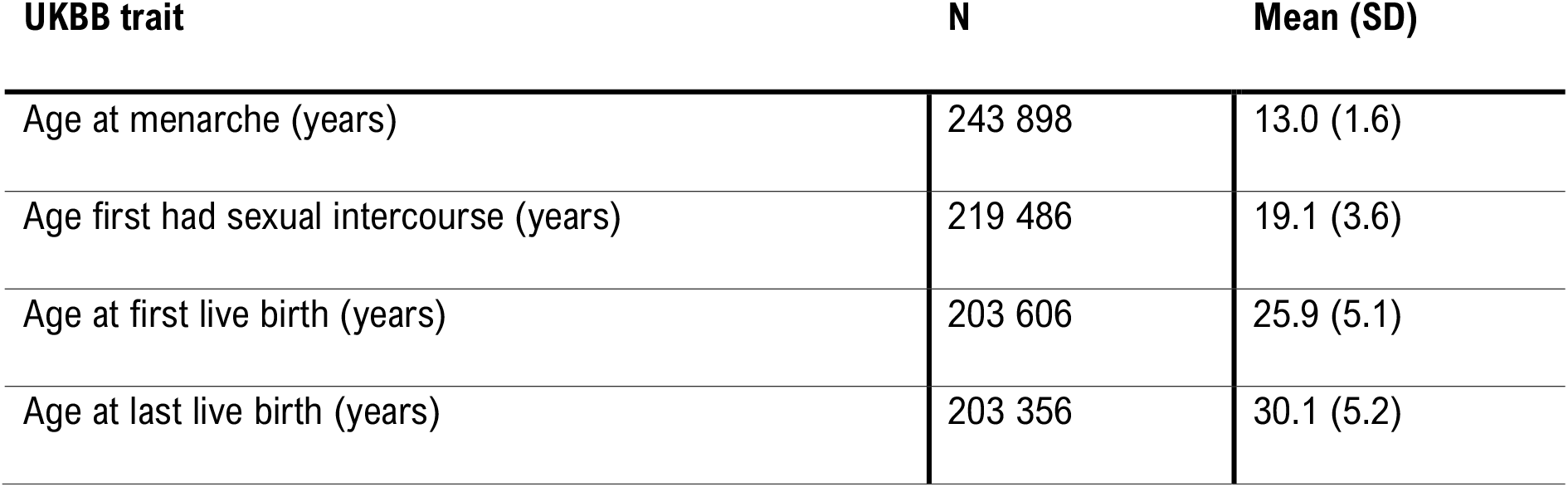
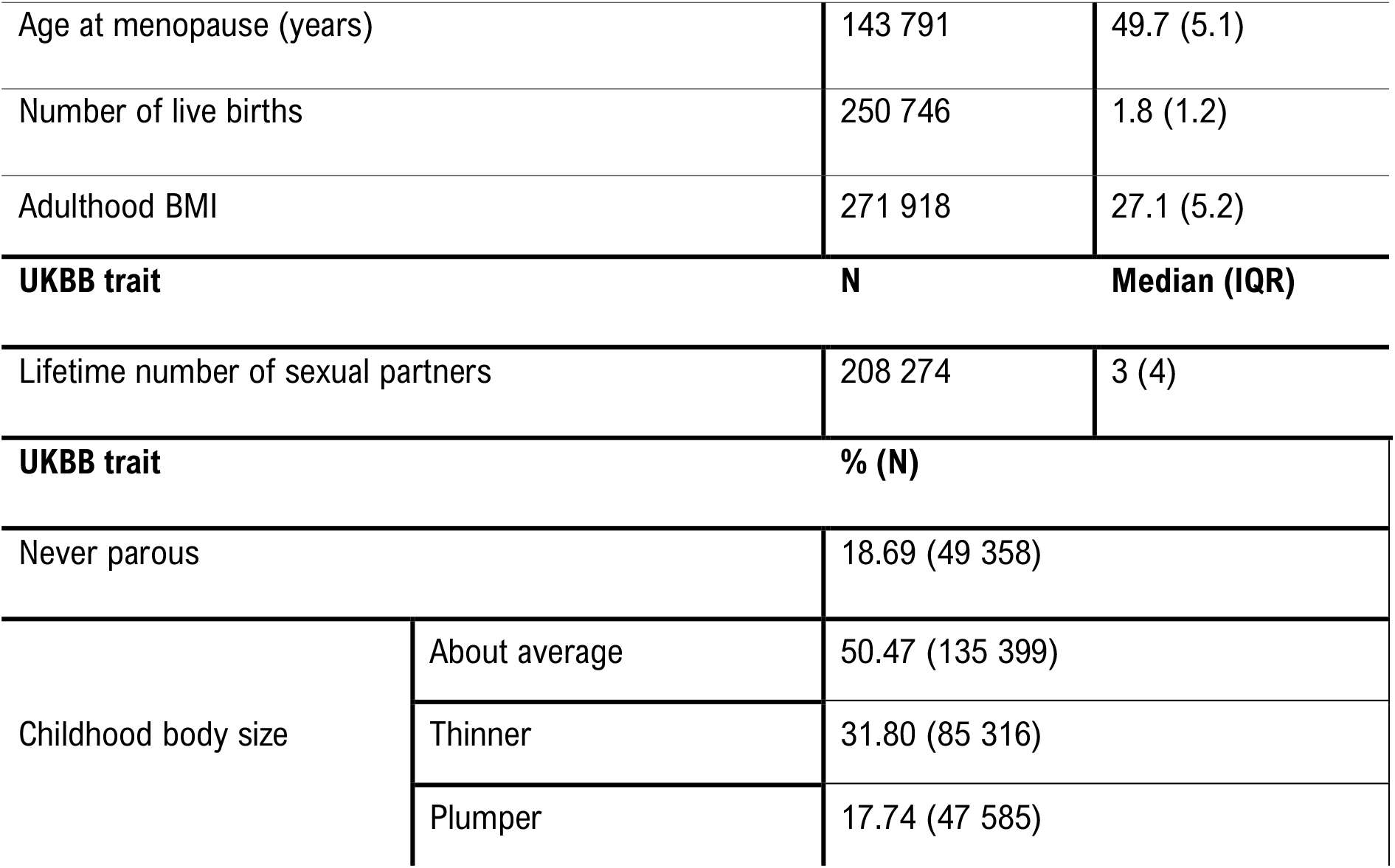
UK Biobank study characteristics. N=Sample size, SD= Standard deviation. IQR = Interquartile range.

| UKBB trait | N | Mean (SD) |
| --- | --- | --- |
| Age at menarche (years) | 243 898 | 13.0 (1.6) |
| Age first had sexual intercourse (years) | 219 486 | 19.1 (3.6) |
| Age at first live birth (years) | 203 606 | 25.9 (5.1) |
| Age at last live birth (years) | 203 356 | 30.1 (5.2) |
| Age at menopause (years) | 143 791 | 49.7 (5.1) |
| Number of live births | 250 746 | 1.8 (1.2) |
| Adulthood BMI | 271 918 | 27.1 (5.2) |
| **UKBB trait** | **N** | **Median (IQR)** |
| Lifetime number of sexual partners | 208 274 | 3 (4) |
| **UKBB trait** | **% (N)** |  |
| Never parous | 18.69 (49 358) |  |
| Childhood body size | About average | 50.47 (135 399) |
|  | Thinner | 31.80 (85 316) |
|  | Plumper | 17.74 (47 585) |

### UK Biobank GWAS

Table 2 displays the number of SNPs associated with each reproductive factor and adiposity measures at genome-wide significance (p value<5×10^−8^) after linkage disequilibrium (LD) clumping within the full UK Biobank sample. In the univariable analysis, all traits had a F statistic over the standard threshold of 10 (Table 2). However, in the multivariable analysis the conditional F statistic was reduced for all traits (Table S1) and was below 10 for AFS, AFB, age at last birth (ALB), number of births, ever parous status and lifetime number of sexual partners with adjustment for other reproductive factors in the MVMR analysis (Table S4).

**Table 2.** Instrument strength of each trait of interest.

| Trait | N | nSNPs | R<sup>2</sup> | F statistic |
| --- | --- | --- | --- | --- |
| Age at menarche | 243 898 | 223 | $6.42 \times 10^{-2}$ | 74.95 |
| Age first had sexual intercourse | 219 486 | 53 | $9.44 \times 10^{-3}$ | 39.46 |
| Age at first live birth | 203 606 | 41 | $8.38 \times 10^{-3}$ | 41.97 |
| Age at last live birth | 203 356 | 9 | $1.92 \times 10^{-3}$ | 43.46 |
| Number of live births | 250 746 | 9 | $1.68 \times 10^{-3}$ | 46.75 |
| Ever parous status | 250 746 | 4 | $8.59 \times 10^{-4}$ | 53.92 |
| Age at menopause | 143 791 | 84 | $4.74 \times 10^{-2}$ | 85.08 |
| Lifetime number of sexual partners | 208 274 | 34 | $6.36 \times 10^{-3}$ | 39.20 |
| Childhood body size | 250 746 | 112 | $3.74 \times 10^{-2}$ | 76.03 |
| Adulthood BMI | 273 382 | 160 | $3.23 \times 10^{-2}$ | 56.95 |

### Mendelian randomization

#### Effects of childhood body size on reproductive factors

Findings referred to here are shown in **Figure 1** and in **Tables S5-S6**. All effects are displayed as per 1 SD increase.

**Figure 1.**
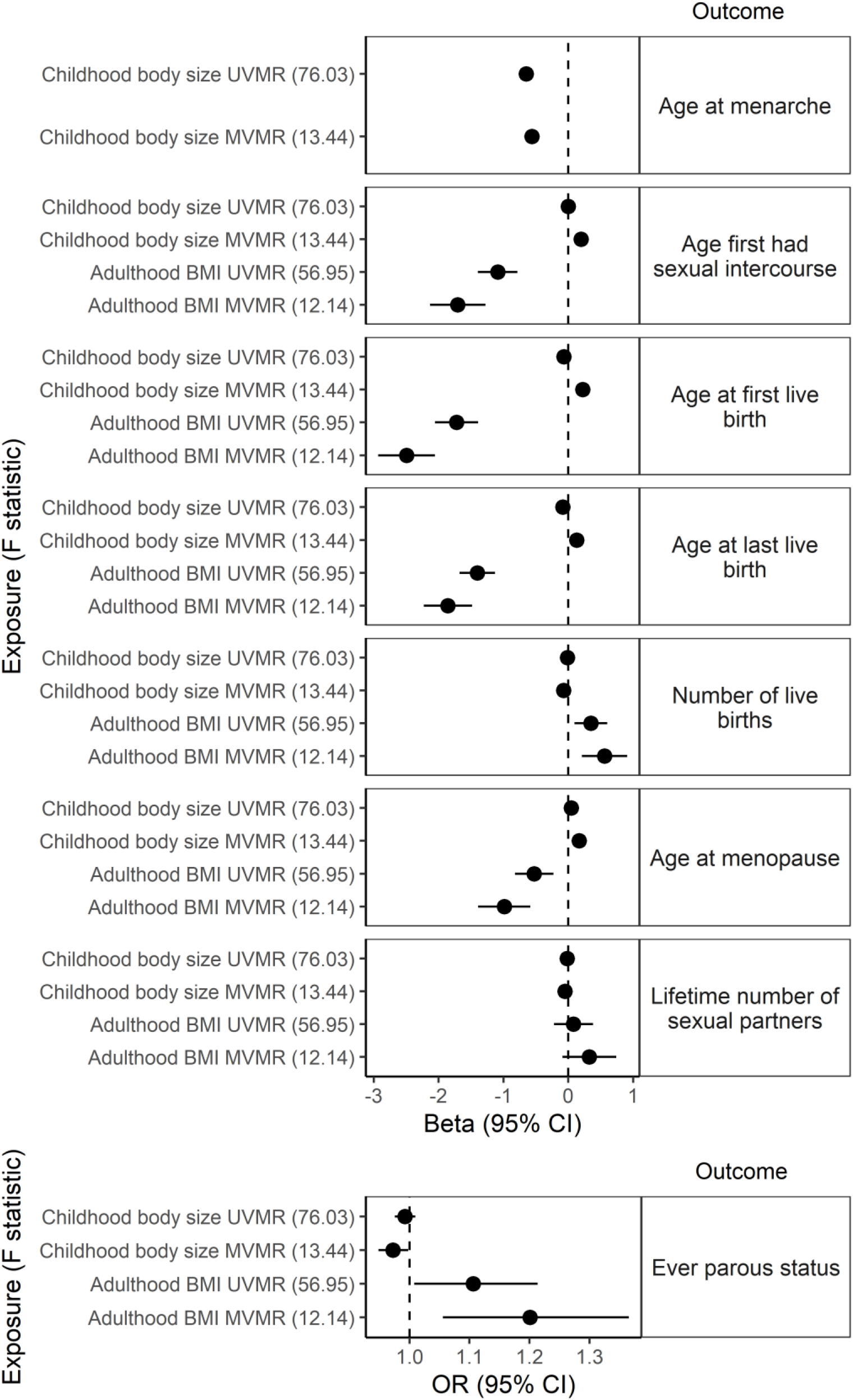
Univariable Mendelian randomization (UVMR) and Multivariable Mendelian randomization (MVMR) findings: the effects of childhood body size (MVMR adjusted for adulthood BM I) and adulthood BMI (MVMR adjusted for childhood body size) on reproductive factors in UK Biobank. GWAS regression coefficients were standardised prior to performing MR. CI – Confidence interval.

The UVMR analysis revealed inverse effects of childhood body size on AAM (Beta (B)=-0.65 SD, 95% confidence interval (CI)=-0.74,-0.57), AFB (B=-0.07 SD, CI=-0.12,-0.01), ALB (B=-0.09 SD, CI=-0.13,-0.04), and no evidence for an effect on AFS (B=-0.002 SD, CI=-0.059,0.054) or AMP (B=0.05 SD, CI=-0.01, 0.11). Conversely in the MVMR model, adjusting for adulthood BMI, the effects were positive for AAM (B=-0.56 SD, CI=-0.68,-0.44), AFS (B=0.20 SD, CI=0.11,0.28), AFB (B=0.22 SD, CI=0.14,0.31), ALB (B=0.13 SD, CI=0.06,0.21) and AMP (B=0.17 SD, CI=0.09,0.25).

Findings from the UVMR analysis suggested there is no evidence for an effect of childhood body size on number of births (B=-0.01 SD, CI=-0.06,0.04), or ever parous status (Odds ratio (OR)=0.99, CI=0.98,1.01), however in the MVMR model, adjusting for adulthood BMI, there was evidence for inverse effects on number of births (B=-0.07 SD, CI=-0.14,-1.74×10^−3^) and ever parous status (OR=0.97, CI=0.95,1.00).

There was no evidence for an effect of childhood body size on lifetime number of sexual partners in the UVMR model (B=-0.02 SD, CI=-0.08,0.04) or in the MVMR model adjusting for adulthood BMI (B=-0.05 SD, CI=-0.13, 0.03).

#### Effect of adulthood BMI on reproductive factors

Findings referred to here are shown in Figure 1 and in Tables S5-S6. All effects are displayed as per 1 SD increase.

In the UVMR model we identified inverse effects of adulthood BMI on AFS (B=-1.09 SD, CI=-1.40,-0.782), AFB (B=-1.72 SD, CI=-2.06,-1.39), ALB (B=-1.40 SD, CI=-1.68,-1.13), and AMP (B=-0.53 SD, CI=-0.82,-0.23). These effects were maintained or strengthened in the MVMR analysis adjusting for childhood body size; AFS (B=-1.70 SD, CI=-2.13,-1.28), AFB (B=-2.48 SD, CI=-2.93,-2.06), ALB (B=-1.86 SD, CI=-2.23,-1.48) and AMP (B=-0.99 SD, CI=-1.39,-0.59).

The UVMR analysis revealed evidence for a positive effect of adulthood BMI on number of births (B=0.35 SD, CI=0.10,0.60) and ever parous status (OR=1.11, CI=1.01,1.21) which were maintained in the MVMR model adjusting for childhood body size (B=0.56 SD, CI=0.21,0.91 and OR=1.20, CI=1.06,1.37).

There was no evidence for an effect of adulthood BMI on lifetime number of sexual partners in both the UVMR model (B=0.08 SD, CI=-0.22,0.38) and MVMR model after adjusting for childhood body size (B=0.33 SD, CI=-0.09,0.74).

#### Effect of reproductive factors on adulthood BMI

Findings are displayed in Figure 2 and in Tables S5-6. All effects are displayed as per 1 SD increase.

**Figure 2.**
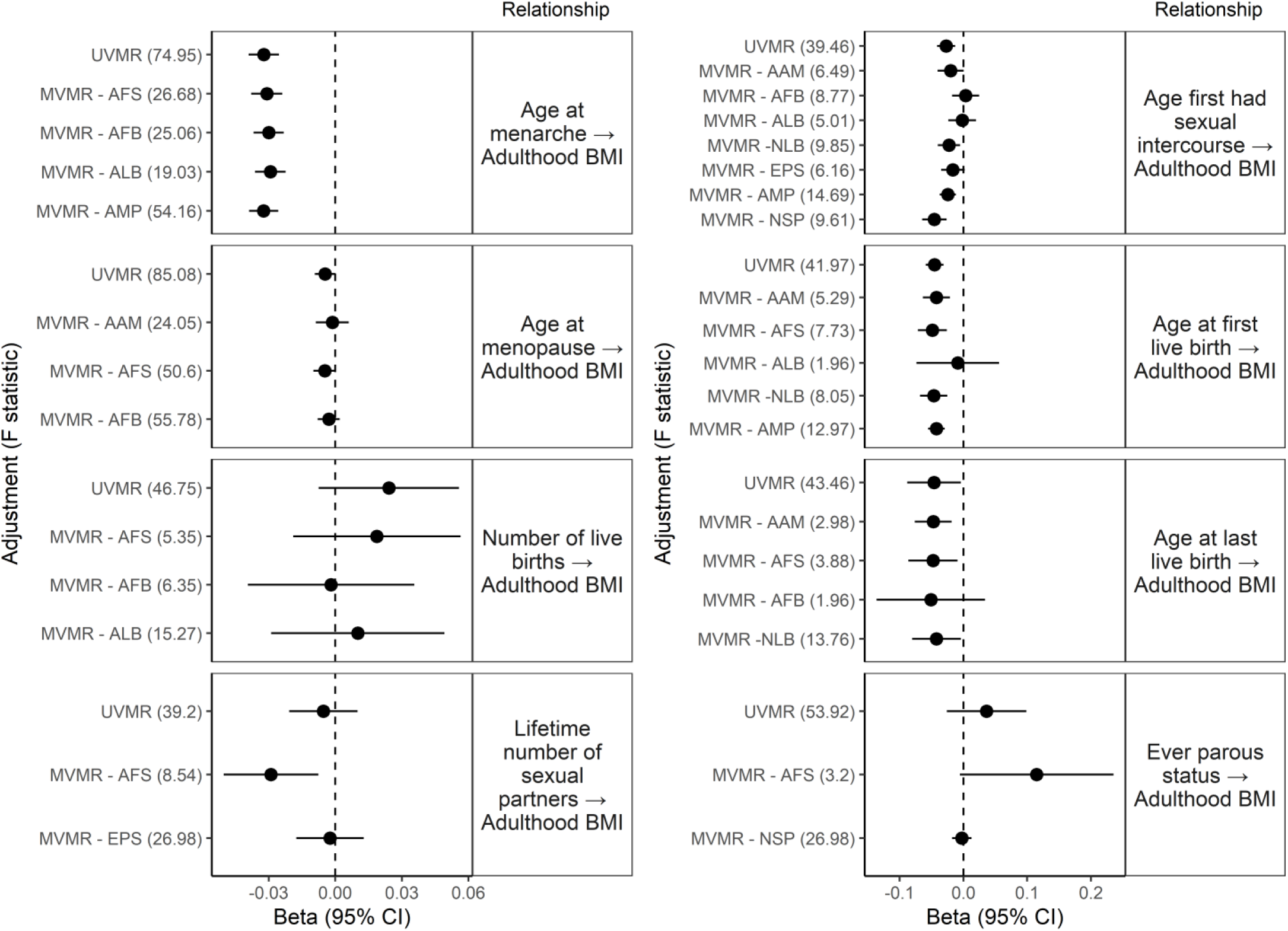
UVMR and MVMR findings: the effects of reproductive factors on adulthood BMI in UK Biobank. GWAS regression coefficients were standardised prior to performing MR. AAM – Age at menarche, AFS – Age at first sexual intercourse, AFB – Age at first live birth, ALB – Age at last live birth, NLB – Number of live births, EPS – Ever parous status, AMP – Age at menopause, NSP – Lifetime number of sexual partners. CI – Confidence interval.

The UVMR analysis of AAM on adulthood BMI revealed evidence for a small inverse effect (B=-3.22×10^−2^ SD, CI=-3.90×10^−2^,-2.53×10^−2^) which did not change with adjustment for relevant reproductive factors in the MVMR. We also found small inverse effects of AFS (B=-2.70×10^−2^ SD, CI=-4.12×10^−2^,-1.29×10^−2^), AFB (B=-4.52×10^−2^ SD, CI=-5.91×10^−2^,-3.13×10^−2^), ALB (B=-4.59×10^−2^ SD, CI=-8.80×10^−2^,-3.90×10^−3^) and AMP (B=-4.62×10^−3^ SD, CI=-9.22×10^−3^,-2.59×10^−5^) on adulthood BMI. However, these effects completely attenuated after adjustment for the relevant reproductive factors in the MVMR models. While we did not find evidence for an effect of lifetime number of sexual partners in the UVMR model (B=-5×10^−3^, CI=-0.02,0.01), adjustment for AFS revealed a small inverse effect (B=-0.03 SD, CI=-0.05,-7.64×10^−3^)

### Evaluating Mendelian assumptions

Details on findings of analyses evaluating MR assumptions can be found in the supplementary note and Tables S7-15.

#### Univariable MR

While the MR Egger, weighted median and weighted mode MR methods where inconsistent with the IVW method (Table S9), suggesting evidence of pleiotropy, the MR PRESSO method revealed little change in strength of evidence after adjustment for outliers (Table S11). MRlap findings suggests bias may have arisen due to sample overlap, and that the effect of adulthood BMI on reproductive factors may be overestimated by ∼19% (range: 15-28%) while the effect of each reproductive factor on adulthood BMI may be underestimated by ∼12% (range: 8-17%) **(Table S12)**.

#### Multivariable MR

For the most part, instrument strength in MVMR is greatly reduced compared to the UVMR analysis. Since evidence of heterogeneity across all relationships, MVMR with minimised Q-statistic was performed which allows for heterogeneity and weak instruments. This revealed similar strength of evidence across most MVMR analyses. Of note the effect of age at menopause on adulthood BMI (adjusting for age first had sexual intercourse) revealed evidence for a very small inverse effect (B=-4.07×10^−3^ SD, CI=-6.74 ×10^−3^, -5.78 ×10^−4^ per 1 SD increase).

### Replication analyses

We performed the UVMR and MVMR analysis using additional non-UK Biobank GWAS summary statistics where possible. Table 3 and Table S16 display the number of SNPs associated the replication with each reproductive factor and adiposity measures, at genome-wide significance (p<5×10^−8^) after LD clumping, for the UVMR and MVMR analyses respectively.

**Table 3.** Instrument strength of each replication trait of interest. N=Sample size, nSNPs= Number of SNPs at genome-wide significance (p<5×10^−8^) after LD clumping.

| Trait | N | nSNPs | R<sup>2</sup> | F statistic |
| --- | --- | --- | --- | --- |
| Childhood BMI (from EGG) | 39618 | 16 | 0.0236 | 59.89 |
| Adulthood BMI (from GIANT) | 171970 | 37 | 0.0145 | 68.25 |
| Age at menarche (from ReproGen) | 182416 | 60 | 0.0261 | 81.52 |
| Age at menopause (from ReproGen) | 69360 | 39 | 0.0420 | 77.89 |
| Age at first birth (from SSGAC) | 189656 | 5 | $9.14 \times 10^{-4}$ | 34.70 |

Effects identified in the UVMR analyses were replicated except for the effect of childhood body size on AFB, adulthood BMI on AFS and ever parous status, and effects of ALB and AMP on adulthood BMI. (Table S17)

Effect identified in the MVMR analyses were replicated except for the effects of childhood body size on AFS, AFB, ALB, ever parous status, and AMP adjusted for adulthood BMI. Additionally, effects of adulthood BMI on AFS, ALB and AMP were not replicated. Furthermore, the effect identified in the MVMR analysis of lifetime number of sexual partners on adulthood BMI after adjustment for AFS was not replicated. (Table S18)

It is worth noting that the UVMR model of adulthood BMI on number of births and AMP, (Table S17), and MVMR model of childhood body size on number of births (adjusted for adulthood BMI) and adulthood BMI on AFB and number of births (adjusted for childhood body size) (Table S18), were only replicated were the replication GWAS summary statistics were used for the outcome.

## Discussion

In this study we used both UVMR and MVMR to investigate the causal relationships between childhood and adulthood adiposity measures and reproductive factors.

After accounting for the genetic correlation between childhood body size and adulthood BMI, and between reproductive factors, the MVMR findings show evidence that higher childhood body size leads to an earlier AAM, and earlier AAM leads to higher adulthood BMI. In addition, the MVMR analysis revealed that higher childhood body size leads to experiencing first sexual intercourse at a later age, later AFB and ALB, older AMP, and not having children. Furthermore, using MVMR we show a higher adulthood BMI appears to lead to an earlier AFB, ALB and AMP, and increased the likelihood of ever having children and having a higher number of children.

It is worth highlighting that across all reproductive factors, the effects of childhood body size and adulthood BMI act in opposite directions in the MVMR analysis suggesting adiposity in earlier life may affect reproductive factors through different mechanisms compared to later in life.

The effect of childhood body size on AAM and number of births (adjusted for adulthood BMI), adulthood BMI on AFB and number of births (adjusted for childhood body size), and AAM on adulthood BMI (adjusted for relevant reproductive factors) were replicated when we used genetic data from additional datasets in the MVMR models. However, the other relationships identified in the MVMR primary analysis were not replicated. In almost all instances this may be due to weak instruments. Weak instruments in the replication MVMR analysis were a particular concern for analyses of the effects of reproductive factors on adulthood BMI, adjusting for other relevant reproductive factors. For example, previous work has revealed that AFB and ALB are highly genetically correlated (rg=0.94, p<0.001) and causally linked (IVW B=0.72, CI=0.67, 0.77).^(15)^ Therefore, in the MVMR the instruments for both factors are weak when adjusting for the other, making it difficult to isolate the direct effect of AFB and ALB.

In analyses of childhood body size and adulthood BMI on AMP, instruments were strong, but the relationships were not replicated. This suggests overestimation of effects in the primary MVMR analysis may be due to bias arising from winner’s curse,^(28,29)^ or sample overlap between the exposure and outcome GWAS populations.^(30)^

We identified an inverse effect of adulthood BMI on AFS in both the UVMR model and the MVMR model adjusting for childhood body size. Given that first experience sexual intercourse is commonly experienced prior to adulthood (In UK Biobank, 64% of women had sexual intercourse before the age of 20), we assume that the genetic variants identified in relation to adulthood BMI in UK Biobank, where participants were assessed between the ages of 40 and 70 years, are stable across adulthood and are therefore are valid instruments for early adulthood adiposity. This is a similar assumption that was made in a recent study which assessed childhood and adulthood adiposity in relation to smoking initiation (mean age 17.8 years).^(31)^ In support of this, an evaluation of the HUNT study identified the crossover of variance explained by childhood to adulthood adiposity genetic scores occurs in late adolescence/early adulthood,^(17)^ suggesting that the genetic liability of adiposity from childhood to adulthood changes during puberty and is stable thereafter. Nevertheless, it may be useful to consider a third adiposity measure in relation to AFS, using a GWAS performed in late adolescence or early adulthood which, to the best of our knowledge, is not currently available.

Related to this, is the issue that it may seem implausible to adjust for adulthood BMI in the MVMR analysis between childhood body size and age at menarche, since adulthood BMI, measured between age 40 and 70 years in UK Biobank, can neither act as a mediate or confounder due to the temporal ordering. However, it is still important to ensure that the estimates for the direct effect of childhood body size are not influenced by underlying genetic correlation with adulthood adiposity, hence why adulthood BMI was included in the MVMR model. ^(31)^ It is also worth noting that conditioning on adulthood BMI in this relationship may cause collider bias since age at menarche has an effect on adulthood BMI, as highlighted in a recent MVMR study of time-varying exposures.^(31)^ In order to avoid this collider bias, we removed variants, from the childhood body size and adulthood BMI instruments, that were identified by the Steiger test of adulthood BMI on each reproductive factor prior to performing MVMR of childhood body size on each reproductive factor adjusted for adulthood BMI. The results of these analysis revealed no difference to the main MVMR analysis suggesting collider bias may not be an issue here.

### Potential mechanisms

We identified an inverse relationship between childhood body size and AAM, in both the UVMR model and MVMR model adjusting for adulthood BMI, which supports findings from previous observational and MR studies.^(7,13)^ This relationship is likely due to increased production of adipocytokines from adipose tissue which may influence pubertal timing,^(32)^ and increased levels of leptin, in response to increased body weight, which is necessary for pubertal onset.^(33)^

It has been suggested that higher adulthood BMI leads to a later AMP due to the effects of adipose tissue leading to increased oestrogen and other endogenous hormone levels.^(34)^ While we found a higher childhood body size leads to a later AMP, we found that higher adulthood BMI reduced AMP, with an inverse relationship also being observed in replication analyses, although of a smaller magnitude. This may be due to higher BMI depleting ovarian reserves, which causes menopause to occur earlier.^(5)^

We found an inverse relationship between AAM and adulthood BMI, which concurs with previous research.^(1,11,12)^ This is likely due to AAM leading to increased exposure to endogenous hormones such as oestrogen and progesterone which causes accelerated physical and psychological changes.^(35)^

The current study found evidence for positive effects of adulthood BMI on number of births and ever parous status. Previous work has suggested that increased adiposity and obesity contribute to subfertility due to biochemical disruptions which include insulin resistance and adipocyte hyperactivation, which in then may lead to impaired endocrine responses in women including lower synthesis of oestrogens and luteinizing hormone, and a greater production of androgens in women.^(36)^ Furthermore there is evidence that increased BMI can lead to conditions associated with decreased fertility.^(37)^

In addition, low adiposity may contribute to reduced fertility as undernutrition may reduce functioning of the reproductive system and lead to defective concentrations of adipocyte-related regulators of endocrine processes such as leptin.^(38,39)^ Previous research has shown a J shaped association between BMI and subfertility,^(40)^ however it is worth noting that our analysis only evaluated linear effects. Nevertheless, later AFB in UK Biobank may not be related to reduced fertility and rather reflect a choice to have children later in life. This is consistent with our findings of a higher BMI in adulthood leading to an earlier age at first birth. But it is worth noting that this differs to the positive effect we identified of childhood body size on AFB. This suggests there may be opposing biological and social mechanisms in action depending on when higher adiposity is experienced across the lifecourse.

Higher adiposity in childhood, in comparison, delayed AFS, this may be explained by overweight in adolescence resulting in later engagement in sexual activity due to social stigma.^(41-43)^

### Limitations

This study had a number of limitations. Firstly, the childhood BMI GWAS from the EGG consortium used in the replication analyses was not sex specific and had a lower sample size compared to the measure from UK Biobank. The lower sample size reduced instrument strength, and childhood BMI in boys was additionally captured which may have influenced the results. Unfortunately, there was not a sex specific childhood adiposity GWAS available to use for replication analysis. Secondly, we were not able to use the minimised Q statistic, which aims to obtain estimate which are robust to weak instruments and pleiotropy, for MVMR analysis with fewer than four genetic instruments. This is because this statistic does not perform well with this number of variants and consequently is not reliable. Therefore, further work would be required to untangle the effects of AFB, ALB and ever parous status on adulthood BMI. In addition, AFB, AFS, number of births, and ever having children, are bio-social traits. Therefore, findings are not easily generalisable to settings with different social norms to the UK and may be specific to the UK Biobank and not generalisable to more contemporary populations. We found evidence for effects of childhood body size and adulthood BMI on AFB and ALB however sensitivity analysis demonstrated difficulty separating out the effects of AFB and ALB, likely due to very high genetic correlation leading to inconclusive results.

## Conclusion

In summary, we found evidence for direct effects of childhood body size on age at menarche, and of age at menarche on adulthood body size. We additionally identified some evidence for direct effects of childhood body size and adulthood BMI on a number of other reproductive factors. Of note, the effects of childhood body size and adulthood BMI had opposing effects on numerous reproductive factors, including age first had sexual intercourse, age at first birth, age at last birth, number of births, ever parous status, and age at menopause. Our findings demonstrate the importance of considering a lifecourse approach when investigating the inter-relationships between adiposity measures and reproductive events, as well as the use of ‘age specific’ genetic instruments when evaluating lifecourse hypotheses in a Mendelian randomization framework.

## Data Availability

The availability of all data analysed in this study has been referenced throughout the manuscript and supplementary materials.

http://egg-consortium.org/

https://portals.broadinstitute.org/collaboration/giant/index.php/Main_Page

https://www.reprogen.org/data_download.html

https://www.thessgac.org/

## Acknowledgements

This research has been conducted using the UK Biobank Resource under Application Number 6326. We thank the participants and researchers from the UK Biobank who contributed or collected data.

## Notes

### Competing Interest Statement

The authors have declared no competing interest.

### Funding Statement

All authors work in a unit that receives funding from the University of Bristol and the UK Medical Research Council (MRC) (MC_UU_00011/1, MC_UU_00011/5, MC_UU_00011/6). All authors are members of the Menarche, Menstruation, Menopause and Mental Health (4M) consortium, which is supported by the GW4 Alliance. C.P. is supported by a Wellcome Trust PhD studentship in Molecular, Genetic and Lifecourse Epidemiology (108902/B/15/Z). G.C.S. is supported by the UK MRC (New Investigator Research Grant, MR/S009310/1) and the European Joint Programming Initiative 'A Healthy Diet for a Healthy Life' (JPI HDHL, NutriPROGRAM project, UK MRC MR/S036520/1). L.D.H. is supported by Career Development Awards from the UK MRC (MR/M020894/1). R.C.R. is supported by the CRUK-funded Integrative Cancer Epidemiology Programme (C18281/A19169).

### Author Declarations

UK Biobank received ethical approval from the North West Multi-Centre Research Ethics Committee (REC reference: 16/NW/0274) and was conducted in accordance with the principles of the Declaration of Helsinki.

