## Supplementary Note for "Establishing the relationships between adiposity and reproductive factors: a multivariable Mendelian randomization analysis"

**Methods**

### GWAS

BOLT-LMM was used to conduct the analysis in the GWAS pipeline,^(1)^ which accounts for population stratification and relatedness using linear mixed modelling. Genotyping chip and age were included as covariates. Genome-wide significant SNPs were selected at p <5×10^−8^ and were clumped to ensure independence at linkage disequilibrium (LD) r^2^ < 0.001 and a distance of 10 000 kb using the TwoSampleMR package.^(2)^

We obtained GWAS summary statistics for childhood body size and adulthood BMI from Richardson et al. 2020, ^(3)^ here they performed a GWAS similarly using BOLT-LMM accounting for population stratification and relatedness and including age at baseline and genotyping array as covariates.

### Univariable mendelian randomization

The IVW MR method combines Wald ratios, calculated by dividing the SNP-outcome association by the SNP-exposure association, in a multiplicative random effect meta-analysis where the weight of each ratio is the inverse of the variance of the SNP-outcome association.^(4)^

This method makes a number of assumptions: that the genetic instruments are strongly associated with the exposure; do not share common causes with the outcome; and are not pleiotropic i.e., do not have an effect on the outcome through a pathway other than via the exposure. ^(4)^

### Multivariable mendelian randomization

Multivariable MR (MVMR) is an extension of MR, and the assumptions are also extended. These assumptions are that the genetic instruments must be strongly associated with each exposure given the other exposures included in the model; do not share any common cause with the outcome; and are independent of the outcome given all of the exposures. ^(5,6)^

### Evaluating univariable mendelian randomization assumptions

To evaluate the strength of the genetic instruments, we determined the mean F statistic for each trait, which is calculated based on the variance explained (r^2^) by the genetic instrument and sample size of the exposure.^(7)^

In addition, we assessed evidence for heterogeneity and invalid instruments, which can imply presence of pleiotropy.^(8,9)^ We evaluated this calculating the Cochran's Q statistic between instruments using the TwoSampleMR package.

To evaluate whether the genetic instruments are pleiotropic we performed MR using additional methods: Weighted mode, ^(10)^ Weighted median,^(11)^ and MR Egger. ^(12,13)^ The intercept and 95% confidence interval of the MR-Egger regression line was used to determine directional pleiotropy using the TwoSampleMR package.^(12)^

We also applied the R function MR-PRESSO (Mendelian Randomisation Pleiotropy RESidual Sum and Outlier) to identify and correct for potential outliers (p <0.05).^(14)^

We performed the MR Steiger test and Steiger filtering bi-directionally for the relationships between reproductive factors and adulthood BMI where we found evidence for bidirectional relationships.^(15)^ This was performed to assess whether the hypothesized causal directional of the relationship was correct for each genetic instrument.^(15)^

### Evaluating multivariable Mendelian randomization assumptions

We evaluated the joint instrument strength for the two exposures in the MVMR setting using the Sanderson–Windmeijer conditional F-statistic, ^(16)^ this was calculated using the ‘strength_mvmr()’ function from the “MVMR” R package. ^(17)^

To evaluate evidence of horizontal pleiotropy we used a modified form of Cochran’s Q statistic available from the ‘pleiotropy_mvmr()’ function from the “MVMR” R package. ^(17)^

Where we identify weak instruments and/or evidence of pleiotropy we additionally performed MVMR with minimized Q-statistic allowing for heterogeneity, using the ‘qhet_mvmr()’ function from the “MVMR” R package. ^(17)^

Conditioning on adulthood BMI in the assessment of the effect of childhood body size on reproductive factors that may have an effect on adulthood BMI can cause collider bias. ^(18)^ To avoid this collider bias we first performed the MR Steiger test between adulthood BMI and each reproductive factor to identify SNPs that explain more variation on the reproductive factor than adulthood BMI .^(15)^ We then performed the MVMR analysis investigating the effects of childhood body size on reproductive adjusting for adulthood BMI excluding those Steiger test identified SNPs.

### Replication analyses

In the primary analysis, we performed two sample MR methods solely in UK Biobank and therefore the exposure and outcome samples fully overlap. Large overlap in the sample(s) used to generate genetic variant-exposure and genetic variant-outcome associations can introduce bias in estimates obtained using two-sample MR methods, which could lead to an overestimation of effects. ^(19)^ However, it has been suggested that applying two-sample MR methods in a single sample may be performed within large studies with minimal bias.^(20)^ Since the GWAS used to identify genetic instruments for MR analysis were also identified in UK Biobank, our analysis is also susceptible to potential winner’s curse, which is the overestimation of the SNP effects on the exposure in a discovery GWAS. ^(21,22)^

Given these concerns, we performed replication analyses using samples independent of UK Biobank to evaluate the robustness of our results. We used childhood BMI GWAS summary statistics from the Early Growth Genetics (EGG) consortium,^(23)^ and adulthood BMI GWAS summary statistics from the GIANT consortium.^(24)^ We used age at menarche,^(25)^ and menopause GWAS summary statistics from the ReproGen consortium,^(26)^ and age at first birth and number of births GWAS summary statistics from the Social Science Genetic Association Consortium (SSGAC).^(27)^ We used the replication datasets as the exposures and UK Biobank as the outcome, and vice versa. Further details on the number of studies and sample sizes used for the replication consortia are shown in **Table S3**. There were no replication GWAS summary statistics available for age first had sexual intercourse, age at last birth, ever parous status, or lifetime number of sexual partners.

**Results**

### Steiger filtering for bidirectional relationships

We applied the MR Steiger method to each univariable MR (UVMR) model of reproductive factors and adulthood BMI, where we found evidence of an effect, to assess whether we had captured the intended causal direction. We evaluated this for the relationship between age first had sexual intercourse, age at first birth, age at last birth and age at menopause in relation to adulthood BMI. Findings show aggregated instruments have successfully captured the intended causal direction in all cases (**Table S7**).

### Evaluating UVMR assumptions

We identified evidence for heterogeneity in the individual SNP effects in the IVW across all UVMR models with the exception of the relationship between ever parous status and adulthood BMI (**Table S8**). We therefore investigated the robustness of results using additional MR methods to account for potential pleiotropy, MR Egger, Weighted median, and Weighted mode. For most relationships, results using these methods were inconsistent with the IVW method, with the exception for the effects of childhood body size on age at menarche, and age at menarche on adulthood BMI, which were consistent across these methods. Results from these methods can be found in **Table S9**.

#### MR-Egger intercept test

For the primary UVMR models, the MR-Egger intercept test revealed evidence for directional pleiotropy in the relationship between childhood body size and age at menarche, adulthood BMI and age first had sexual intercourse, age at first birth, age at last birth, age at menopause and ever parous status (**Table S10).**

#### MR-PRESSO

MR-PRESSO revealed outlier SNPs in all the primary UVMR models, which were likely to be driving the levels of heterogeneity. However, after outlier correction, there was little change in the strength of evidence apart from for models of childhood body size on age at menopause where an inverse effect emerged, and models of number of births and adulthood BMI where evidence for a positive effect emerged with outlier correction (**Table S11**).

### MRlap

MRlap findings show a reduction in the magnitude of effects when assessing the effect of adulthood BMI on reproductive factors. In addition, we show an elevation in the magnitude of effect when assessing the effect of each reproductive factor on adulthood BMI (**Table S12**). In both directions the evidence of effects that were identified in the primary UVMR models, were maintained. This suggests bias has arisen due to sample overlap that acts in the opposite direction depending on which direction we are investigating, i.e., effect of adulthood BMI on reproductive factors, or reproductive factor to BMI.

### Evaluating MVMR assumptions

For all relationships assessed in the MVMR primary analysis, we identified evidence of heterogeneity in the individual SNP in the IVW across all investigated relationships (**Table S13**).

Due to evidence of heterogeneity, we additionally performed MVMR with minimised Q-statistic allowing for heterogeneity. These analyses revealed similar strength of evidence across analyses other than for the relationships between childhood body size and ever parous status (adjusting for adulthood BMI), adulthood BMI and number of births (adjusting for childhood body size), adulthood BMI and ever parous status (adjusting for childhood body size), and age at last birth and adulthood BMI (adjusting for number of births), where effects attenuated. Of note the effect of age at menopause on adulthood BMI (adjusting for age first had sexual intercourse) revealed evidence for a very small inverse effect (B=-4.07x10^-3^ SD, CI=- 6.74 x10^-3^, -5.78 x10^-4^ per 1 SD increase) (**Table S14**).

However, this method does not perform well where the instrument strength is less than or equal to 5. We therefore we did not investigated effects of age at first birth on adulthood BMI (adjusting for age at last birth), age at last birth on adulthood BMI (adjusting for age at menarche, age first had sexual intercourse, and age at first birth) and ever parous status on adulthood BMI (age first had sexual intercourse).

We removed Steiger test identified SNPs from the MVMR analysis assessing the effects of childhood body size on reproductive factors adjusting for adulthood BMI, however there were no Steiger identified SNPs between adulthood BMI and ever parous status. There was no change in evidence compared to the primary MVMR analysis other than the effect of childhood body size on number of births attenuated slightly, with 95% confidence intervals crossing the null. In addition, instrument strength remained similar to the primary MVMR analysis. (**Table S15**)

### Replication analyses

In the univariable analysis all replication traits had a F statistic over the standard threshold of 10, however, in the multivariable analysis the F statistic was reduced to ~3.8 for both childhood body size and adulthood BMI, and to 0.9 for age at first birth (from SSGAC) when adjusted for number of births (from SSGAC) (**Table S16**).
